# A proposed grading system for spinal cord arteriovenous shunts

**DOI:** 10.1101/2023.08.30.23294873

**Authors:** Jia-Xing Yu, Hao-Han Lu, Li-Song Bian, Yue-Shan Feng, Jing-Wei Li, Fan Yang, Gui-Lin Li, Chuan He, Ming Ye, Peng Hu, Li-Yong Sun, Yong-Jie Ma, Jian Ren, Feng Ling, Tao Hong, Hong-Qi Zhang

## Abstract

**Background:** The clinical outcomes of microsurgery for spinal cord arteriovenous shunts (SCAVSs) exhibit fluctuations due to varying patient selection criteria, underscoring the importance of a standardized surgical grading system that can effectively stratify the feasibility of SCAVSs resection.

**Methods:** A cohort of 308 consecutive patients with surgically treated SCAVSs was randomly divided into a modeling group and a validation group. The surgical grading system was developed based on the independent risk factors of incomplete resection identified in the modeling group and subsequently verified in the validation group. The system’s specificity and sensitivity were tested through Receiver Operating Characteristic (ROC) analyses.

**Results:** Multivariate analysis indicated that metameric AVSs (p=0.007), AVSs with maximum length ≥3 cm (p=0.017), embedded AVSs (p=0.032) and anterior sulcal artery supply (p=0.013) were independent risk factors of incomplete resection. Subsequently, each of the four parameters is assigned one point, and the SCAVSs grade is calculated by aggregating all parameter scores. The area under ROC curve (AUC) of modeling group and validation group was 0.856 (95% confidence interval [95% CI], 0.794-0.919) and 0.819 (95% CI, 0.747-0.892) respectively. Across the entire cohort, patients with scores ranging from 0 to 4 exhibited complete resection rates of 88.7%, 66.7%, 30.6%, 4.5% and 6.3%. The corresponding rates of severe treatment-related deterioration were found to be at levels of 6.0%, 12.0%, 12.9%, 31.8% and 25.0 %, respectively. Conclusion: The proposed grading system effectively stratifies the surgical feasibility of SCAVSs based on both the probability of achieving complete resection and the treatment risk. Its simplicity renders it a valuable tool for clinical decision-making, as well as a reference point for evaluating treatment outcomes across different centers and surgical techniques.

## Introduction

Clinical intervention is imperative for spinal cord arteriovenous shunts (SCAVSs) due to their unfavorable natural history^1^. Currently, endovascular embolization stands as the most prevalent treatment option^2^. Although reducing the blood flow or occluding the high-risk components such as aneurysms may reduce the risk of spinal cord impairment, the long-term clinical outcomes of endovascular therapy are limited by its low obliteration rate. In contrast, microsurgery demonstrates a significantly higher rate of lesion obliteration, thereby potentially obviating the clinical risks associated with SCAVSs altogether^1–4^. Therefore, surgical resection should be an optimal option when the treatment risk is judged to be acceptable.

However, due to the rarity and highly heterogeneous angioarchitecture of SCAVSs, patients with these challenging lesions are managed primarily based on individual treatment experiences and expertise. This leads to significant variations in treatment outcomes between different centers^5–7^. Therefore, akin to the Spetzler-Martin score utilized for brain AVMs^8^, a grading system that can objectively assess the complete resection rate and surgical risks is imperative for SCAVSs.

In this study, based on clinical data from the largest multicenter cohort of surgically treated SCAVSs, we present a grading system for stratifying surgical feasibility of SCAVSs.

## Methods

### Patients

We reviewed consecutive patients with SCAVSs who were admitted to Xuanwu Hospital, Beijing Haidian Hospital and Beijing United Family Hospital between January 2007 and June 2022. The study was reviewed and approved by the local ethics committee with waiver of informed consent from patients given its retrospective nature.

Patients were included if: (i) they were diagnosed with SCAVSs; (ii) the intradural AVSs location ranged from the atlas to the tip of the conus medullaris; (iii) they had been surgically treated in our institutes; and (iv) they were followed up for at least six months.

Patients were excluded if: (i) they had concurrent tethered cords, spinal tumors or any other kind of disease that impaired spinal cord function; (ii) they underwent SCAVSs resection in the external hospital; and (iii) patients younger than 1 year old were not eligible because their spinal cord function could not be evaluated objectively.

### Digital subtraction angiography (DSA) evaluation

The angioarchitecture parameters of the lesions were extracted from DSA images by experienced interventional neuroradiologists. AVSs are classified as nidus-type or fistula-type depending on to the existence of the intervening nidus; spinal metameric AVSs (SMAVSs) refers to lesions involving at least one other tissue belonging to the same segment as the diseased spinal cord, including nerve roots, vertebrae, paraspinal muscles, as well as skin, etc^1,9–10^. Lesion depth is classified as embedded or superficial based on whether the medial side of the lesion crossed the midline of the spinal canal or not on both the anteroposterior and lateral view angiograms. Lesion segment is classified into the cranio-cervical cord (C1–C2), the mid-cervical cord (C3–C5), the cervicothoracic cord (C6–T2), the mid-thoracic cord (T3–T9) and the thoracolumbar cord (from T10 to the tip of conus medullaris). The relationship between the major part of each lesion and the spinal cord is defined as ventral, lateral, central and dorsal.

Patients in our institutes would receive a DSA immediately after the surgical procedure. Complete resection is defined as the disappearance of the intradural AVSs based on post-operation DSA, with or without residual paravertebral lesions (If the patient were diagnosed with SMAVSs). The insidious AVSs are characterized by tiny and diffuse intramedullary AVSs that remain undetectable by DSA until the majority of the lesion is occluded.

### Treatment

In this cohort, all the patients underwent microsurgery via a posterior approach. For superficial intramedullary AVSs, myelotomy would be directly performed through the surface that closet to the lesion. For deep located (embedded) lesions, the posterior midline myelotomy or the ADREZotomy would be selected based on the horizontal location and size of the AVSs^11,12^. During the operation, we sharply separated the malformed vessels from the spinal cord tissue and took strict measures to avoid coagulating or suctioning the spinal cord.

Since 2013, we performed SCAVSs surgery in hybrid operating room, so that the intraoperative DSA and methylene blue angiography could be used to enhance our intraoperative understanding of angioarchitecture and identify any residual intramedullary lesions. The detailed procedures of intraoperative angiography were reported in our previous study^13,14^. (Video1)

Intraoperative neurophysiological monitoring including somatosensory evoked potential (SEP) and transcranial motor-evoked potential (MEP) was performed for all operations. In general, a 50% irreversible amplitude reduction of the tcMEP criterion was used to warn the surgeon, while a > 80% criterion was used to stop the operation in order to avoid neurological impairments^14^.

### Clinical evaluation

The observational period was defined as the interval between onset and invasive treatment. Follow-up was planned at discharge, 1 month after discharge, 6 months after discharge and then at yearly intervals, via direct clinical interviews or telephone contact. The follow-up period was defined as the interval between microsurgery and the most recent available follow-up.

Spinal cord function was evaluated according to the modified Aminoff and Logue scale (mALS) ^15^. An increase in mALS of 2:1 point that occurred within 2 weeks after treatment and was sustained for more than 6 months or until death was defined as treatment-related deterioration^1^. Meanwhile, we have further categorized treatment-related deterioration as either mild (an increase in mALS of 1 point) or severe (an increase in mALS of >1 point).

### Statistical analysis

IBM SPSS Statistics 26.0 Statistical software to process and check the level α = 0. 05. The parameters conforming to the normal distribution were expressed by (x ± s), and the parameters of non-normal distribution were expressed by median and quartile [interquartile range (IQR)]. Counting data were expressed by n (%). Differences between groups were tested using Pearson’s chi-square test. Categorical variables were tested by Student’s t-test or Mann-Whitney U-test. Multivariable logistic regression model included variables that were significant at P<0.05 in the univariate analysis. SPSS software was used to randomly divide all patients into two groups, one was modeling group, and the other was validation group. Receiver operating characteristic (ROC) analyses were performed to identity predictive accuracy of the grading system. Area under the ROC curve (AUC) of 0.70 or more is considered a clinically useful predictive model^16^.

All statistical tests were two-sided, and P-values<0.05 were considered statistically significant.

### Data availability

Data collected for the study will be made available upon reasonable request to the corresponding authors.

## Results

### Baseline characteristics

The baseline characteristics of the entire cohort were presented in Table 1. A total of 308 SCAVSs patients were included, among whom 126 patients (40.9%) were female. Based on spinal DSA, nidus-type were identified in 239 patients (77.6%), while fistula-type were found in 69 (22.4%) patients. Out of the included patients, 59 (19.2%) were diagnosed as spinal metameric AVSs. All patients in this study were symptomatic with a median onset age at 28 years [interquartile range (IQR) 20-38 years].

**Table 1.**
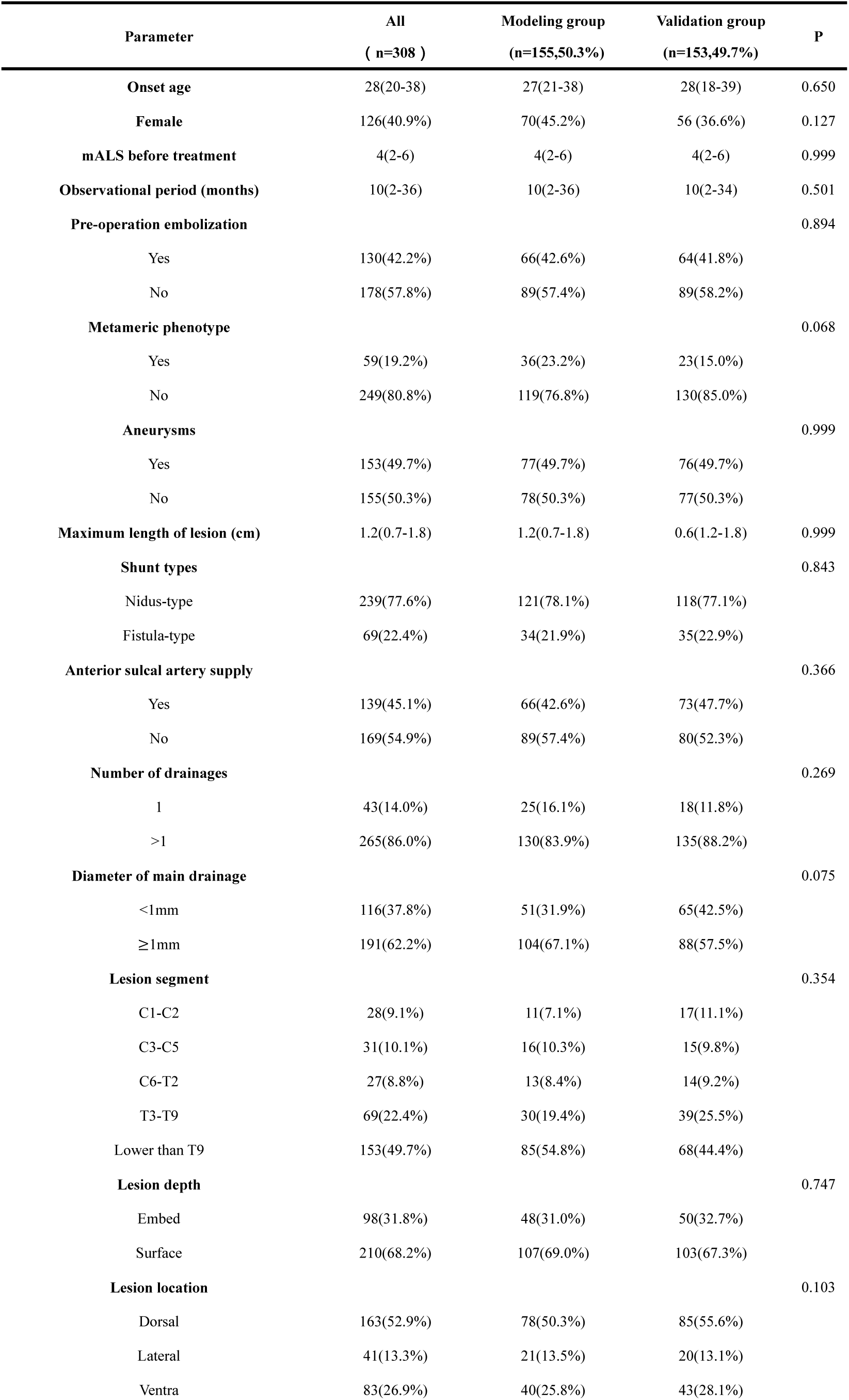
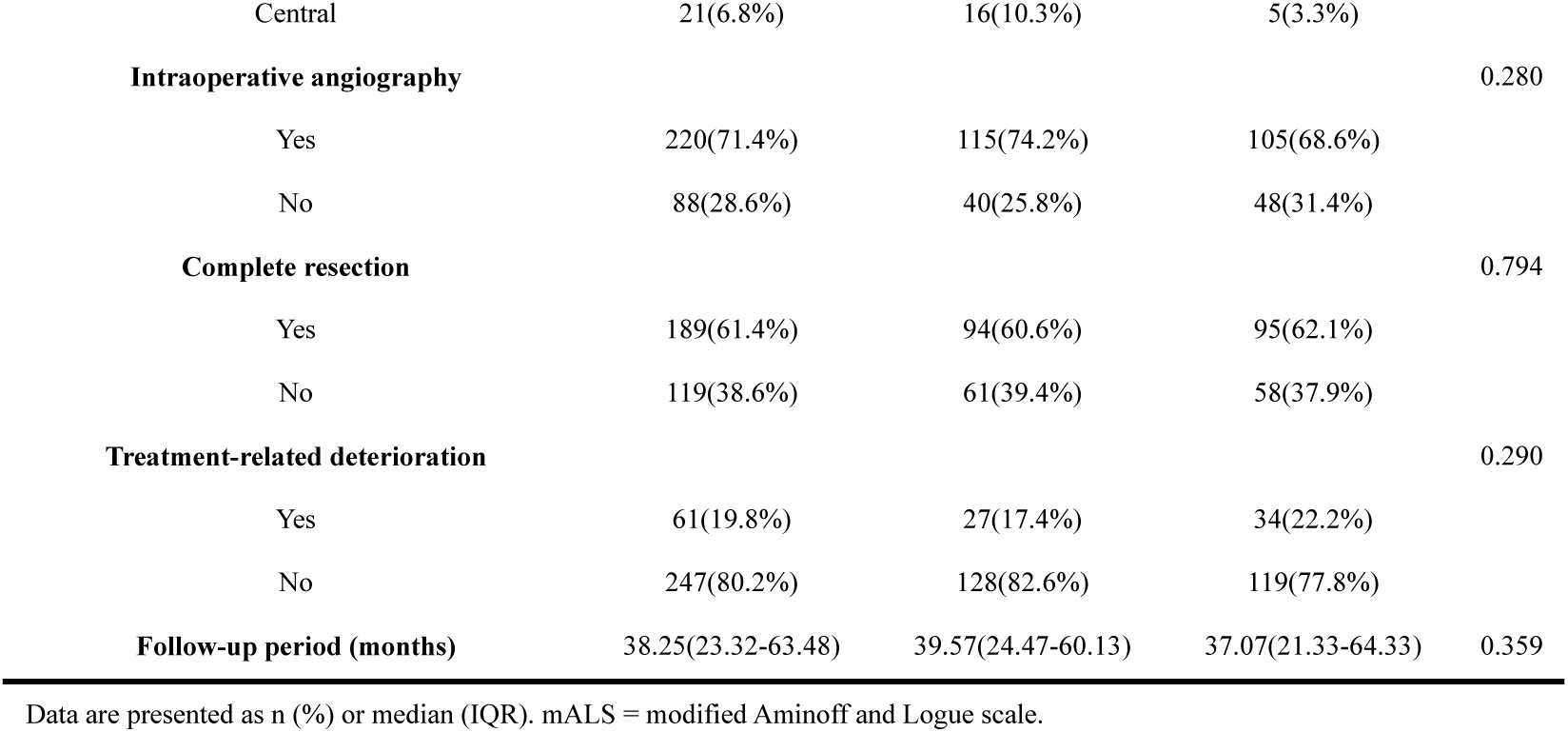
Baseline characteristic.

One hundred and thirty patients (42.2%) underwent preoperative embolization. The overall complete resection rate for the whole cohort was 61.4%. Reasons for failure to complete resection included deliberate partial resection to alleviate symptoms in 28 patients, and interrupted by the intraoperative neurophysiological monitoring in 90 patients (including three patients with insidious intramedullary AVSs) (Figure 1). Sixty-one patients (19.8%) experienced treatment-related deterioration including 25 mild (8.1%) and 36 severe (11.7%) deficits. The media follow-up period of the cohort was 38.25 months (IQR 23.32-63.48 months). At the last available follow-up, 162 patients (52.6%) demonstrated improvement in mALS, while 81 patients (26.3%) remained stable and 4 patients (1.3%) experienced neurological deficits due to residual lesions.

**Figure 1:**
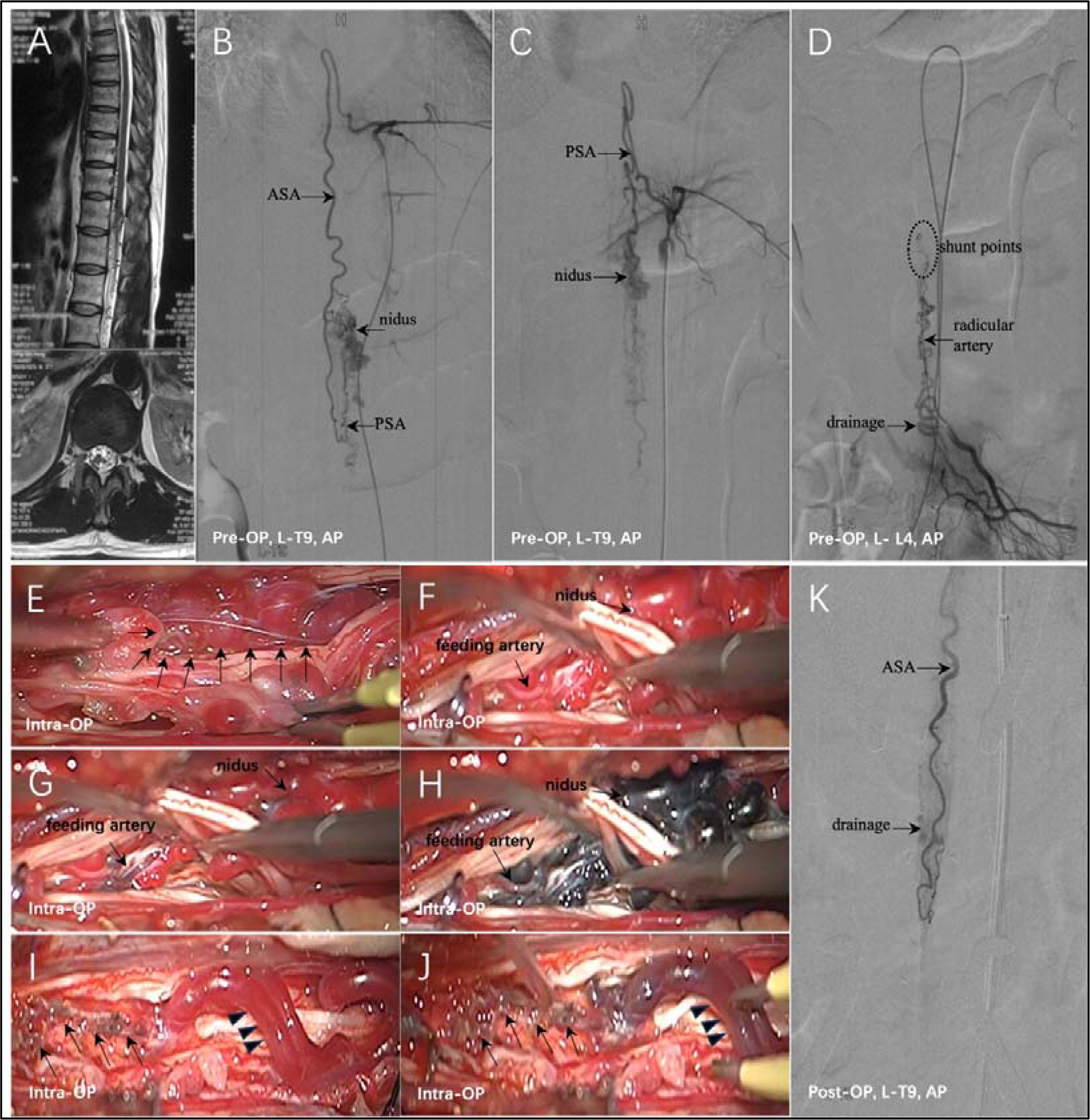
Incomplete resection of a 1-point SCAVSs. This AVSs was a superficial (0 point) metameric lesion (1 point) measuring 2.7cm in length (0 point), lacking anterior sulcal artery supply (0 point). **(A - C)** Pre-operative MR and DSA revealed that the nidus was predominantly located in the left-dorsal region of the conus medullaris. The main feeding artery was left PSA. ASA suppled the shunts though the pial plexus. **(D)** The radicular AVSs that supplied by L-L4 radicular artery indicated the metameric phenotype. **(E)**The patient received surgical treatment in a hybrid operating room. Peri-medullary nidus (arrows) was identified after opening the dura and arachnoid. **(F-H)** Continuous images of methylene blue angiography revealed bluing of feeding artery (pial plexus) and nidus in sequence. **(I-J)** The main parts of the AVSs were resected. However, methylene blue angiography still revealed an early successive bluing of the pial venous network (arrows) on the conus medullaris and dorsal longitudinal veins (arrowheads), indicating the existence of intramedullary AVSs. **(K)** Intra-OP DSA confirmed the intramedullary AVSs. Note the residual AVSs were tiny and scattered in the conus medullaris, making it could not be safely resected. (ASA: anterior spinal artery; PSA: posterior spinal artery; AP: anterior-posterior view; LAT: lateral view; intra-OP: intraoperative; L: left)

### SCAVSs grading scale design

Based on the random numbers generated by SPSS, we had randomly divided all the patients into the modeling group (n=155) and the validating group (n=153). Statistical analysis showed that the clinical characteristics, treatment and clinical outcomes of both groups were well-balanced (Table 1).

To identify potential factors that contribute to the difficulty of SAVS resection, we conducted an analysis of independent predictors of incomplete resection in the modeling group. Univariable logistic regression analysis indicated that metameric phenotype (p<0.001), maximum length of lesion (p<0.001), shunt types (p=0.003), anterior sulcal artery supply (p<0.001), lesion depth (p<0.001) and lesion location (p=0.009) may associate with complete resection rate of the microsurgery, then a multivariate logistic regression analysis indicated metameric AVSs (Odds ratio[OR], 4.087; 95% confidence interval [95% CI], 1.459-11.447; p=0.007), AVSs with maximum length 2:3 cm (OR, 8.598; 95% CI, 1.469-50.319; p=0.017), embedded AVSs (OR, 3.026; 95% CI, 1.101-8.317; p=0.032) and AVSs with anterior sulcal artery supply (OR, 3.396; 95% CI, 1.295-8.906; p=0.013) were independent predictors of incomplete resection. (Table 2)

**Table 2.**
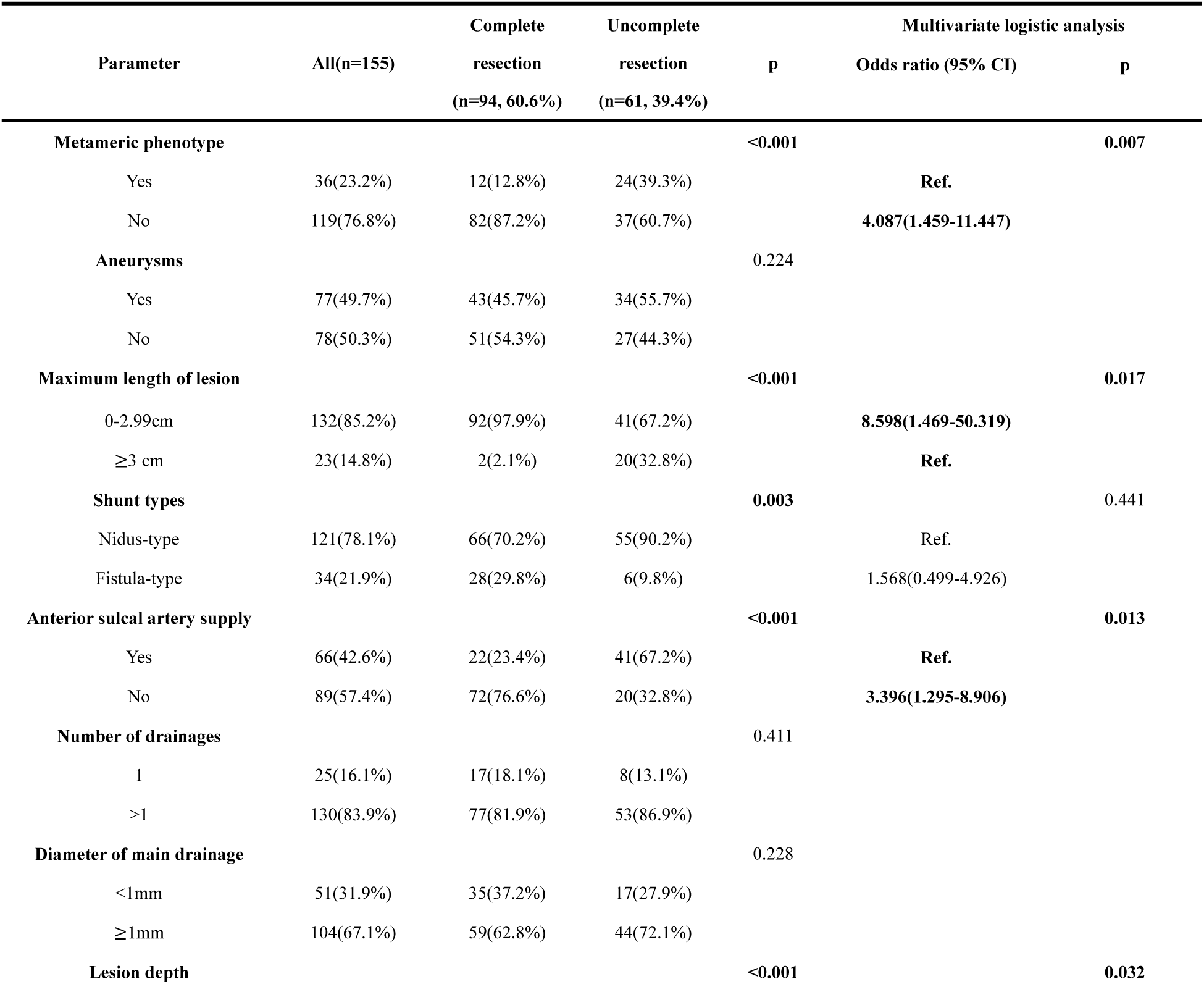
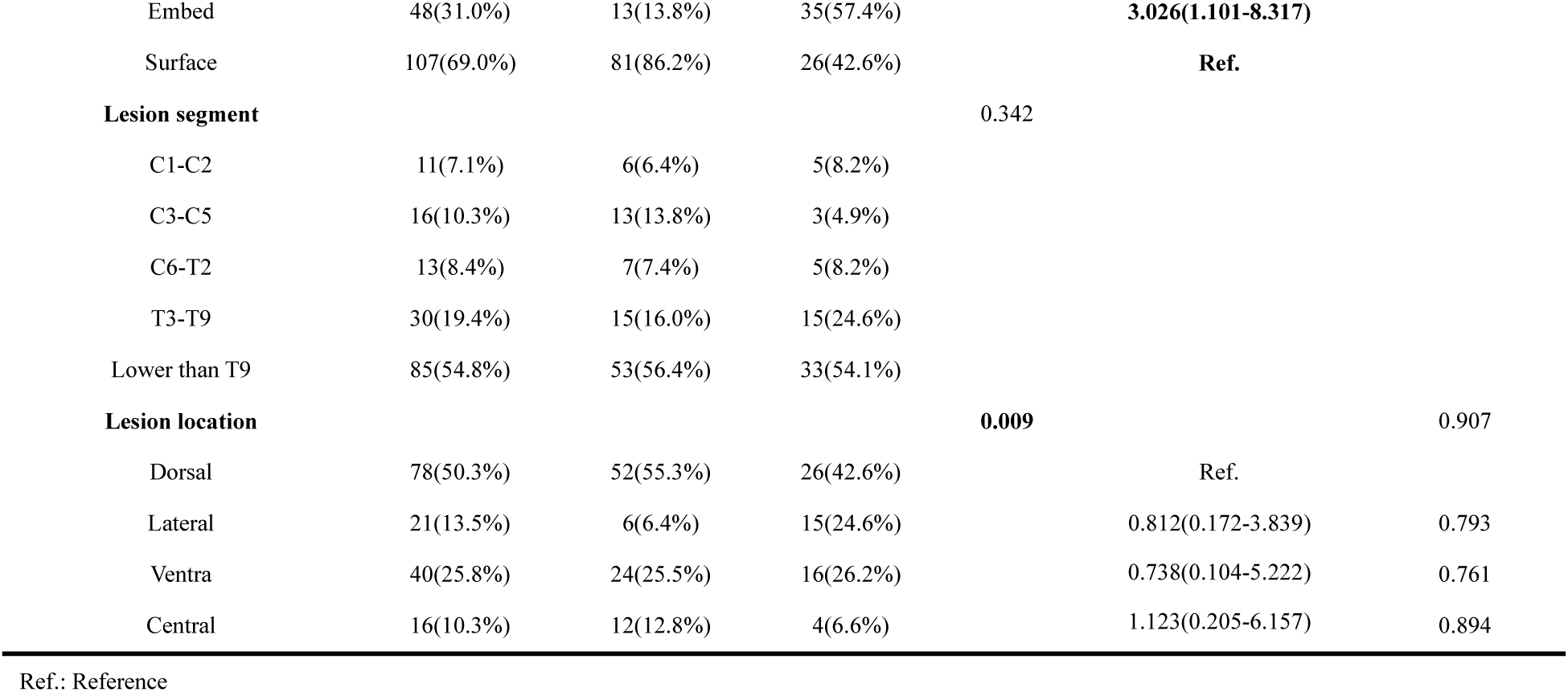
The relationship between angioarchitecture and surgical outcomes in modeling group.

Subsequently, we assigned one point to each of the four factors and calculated the SCAVSs grade by aggregating all parameter scores (Table 3, Figure 2). ROC curve of modeling group showed the AUC of the score was 0.856 (95% CI, 0.794-0.919) (Figure 3 A).

**Figure 2.**
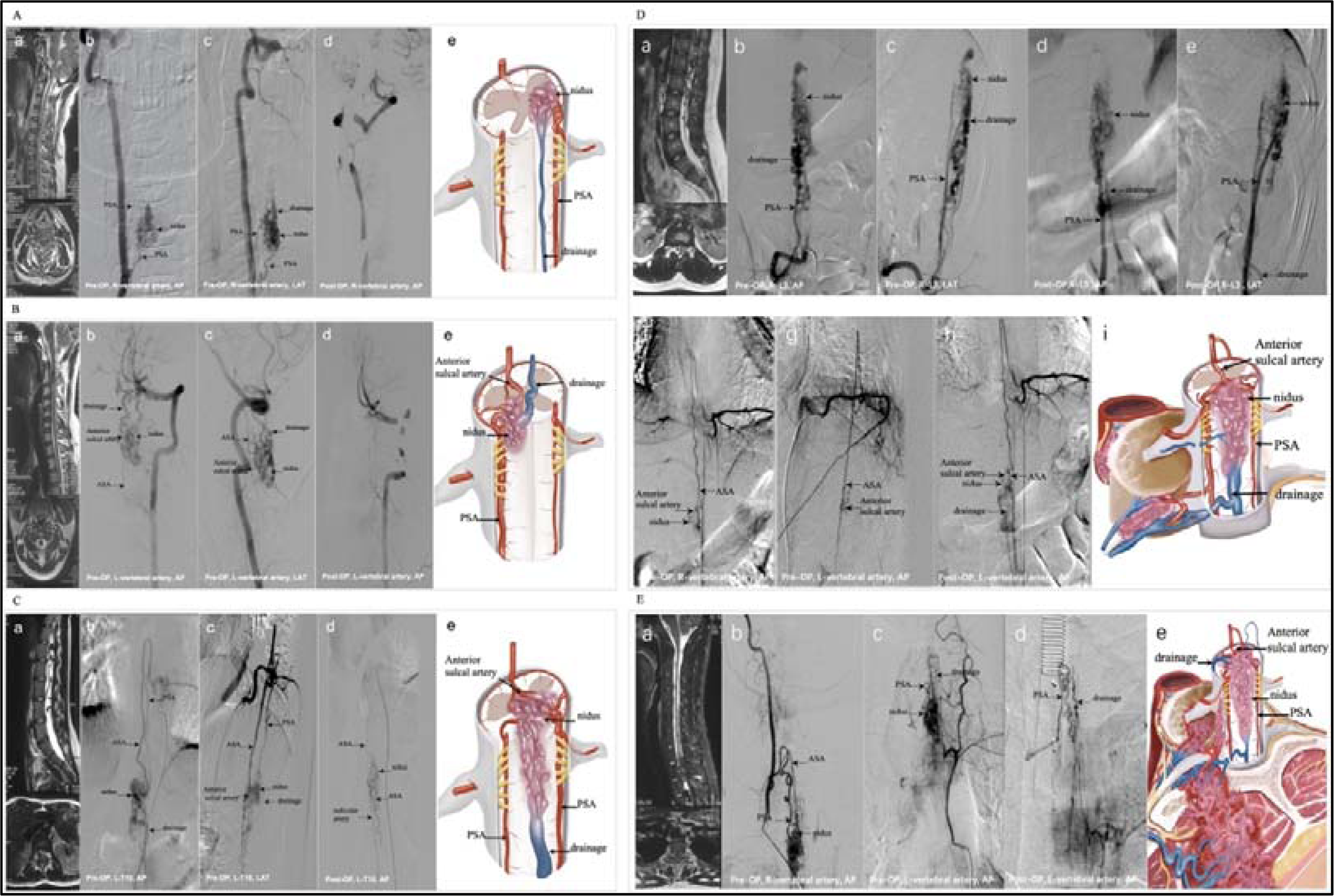
Representative cases scoring between 0 and 4 points according to the SCAVSs surgical grading system. **A.** Complete resection of a 0-point SCAVSs. This AVSs is a superficial (0 point) non-metameric lesion (0 point), measuring 1.2 cm in length (0 point), lacking anterior sulcal artery supply (0 point). **B.** Complete resection of a 1-point SCAVSs. This AVSs is a superficial (0 point) non-metameric lesion (0 point), measuring 1.4 cm in length (0 point) and with anterior sulcal artery supply (1 point). **C.** Incomplete resection of a 2-point SCAVSs. This AVSs is non-metameric lesion (0 point), embedded into the spinal cord (1 point) measuring 2.8cm in length (0 point) and with anterior sulcal artery supply (1 point). **D.** Incomplete resection of a 3-point SCAVSs. This AVSs is a superficial (0 point) metameric lesion (1 point), measuring 4.0 cm in length (1 point) and with anterior sulcal artery supply (1 point). **E.** Incomplete resection of a 4-point SCAVSs. This AVSs is metameric lesion (1 point) embedded into the spinal cord (1 point), measuring 5.4 cm in length (1 point) and with anterior sulcal artery supply (1 point). (ASA: anterior spinal artery; PSA: posterior spinal artery; AP: anterior-posterior view; LAT: lateral view; OP: operation; L: left; R: right)

**Figure 3.**
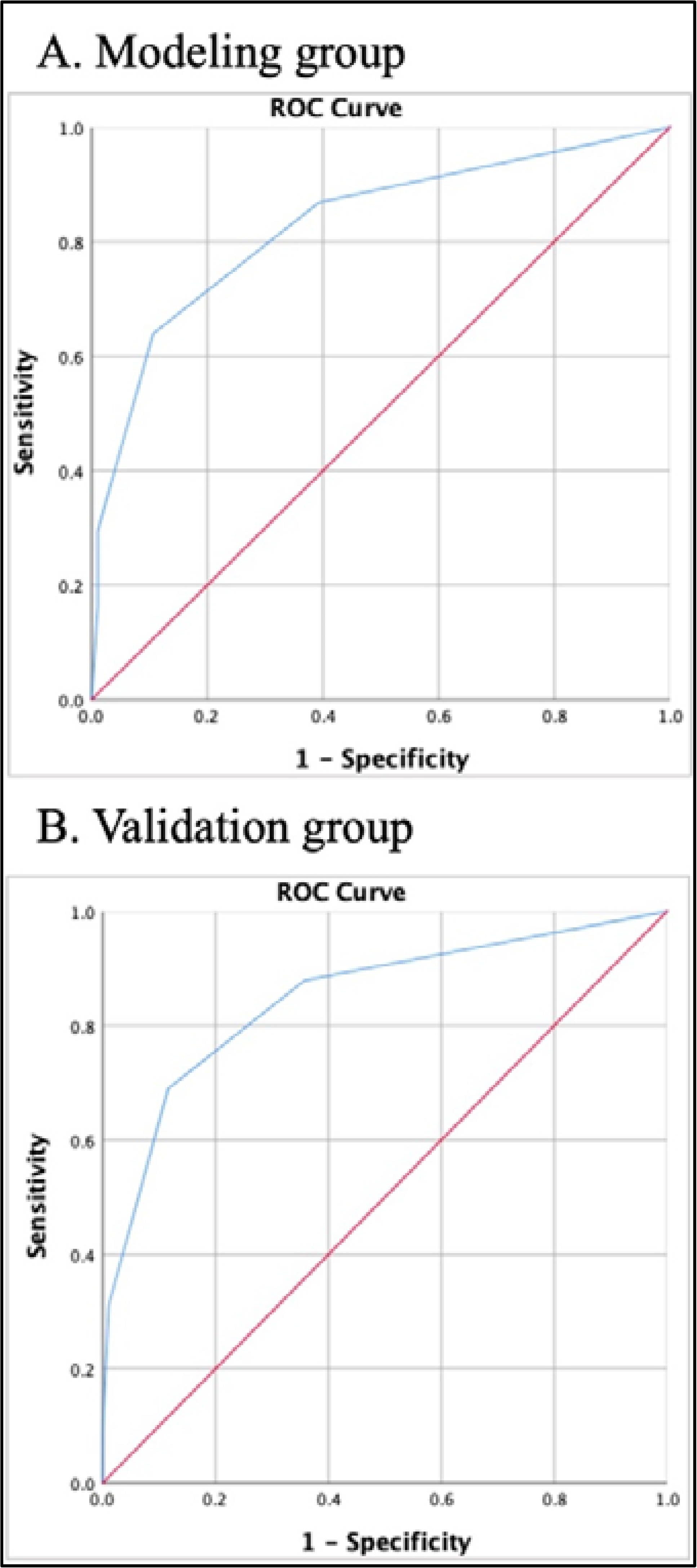
ROC curve of predicting the complete resection rate of SCAVSs operation.

**Table 3.**
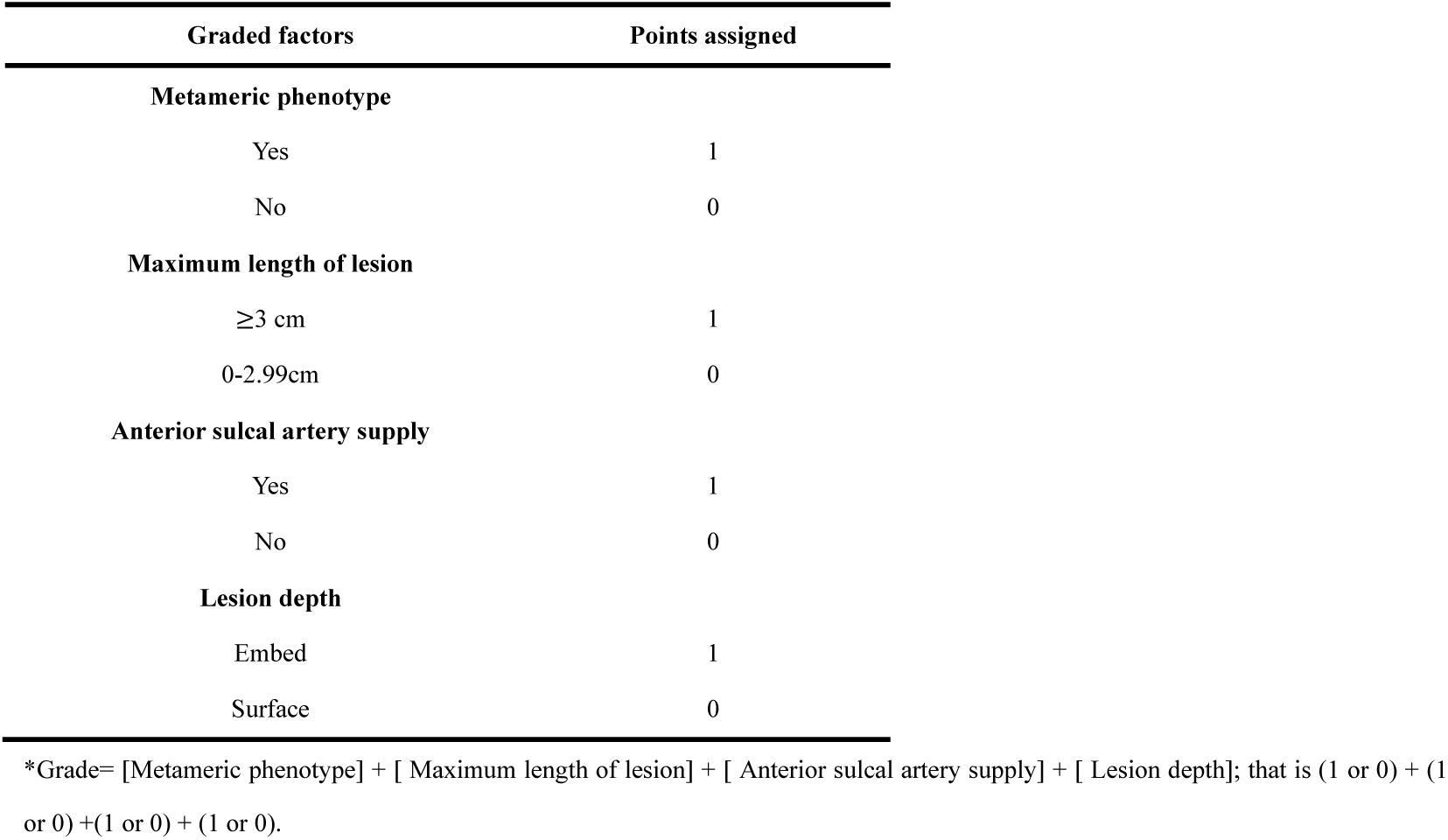
SCAVSs surgery grading system*.

In the validation group, the AUC of ROC curve was 0.819 (95% CI, 0.747-0.892), indicating that the proposed scale exhibited a favorable predictive ability for assessing surgical difficulty in spinal cord AVSs (Figure 3 B).

### The proposed scale stratified surgical outcomes

The entire cohort consisted of 133 patients with a score of 0, 75 patients with a score of 1, 62 patients with a score of 2, 22 patients with a score of 3, and finally, there were 16 patients with a score of 4.

The complete resection rates for scores ranging from 0 to 4 were as follows: 88.7%, 66.7%, 30.6%, 4.5% and 6.3%. (Figure 4A). The incidence of treatment-related deterioration, as measured by scores ranging from 0 to 4, was observed at rates of 18.0%, 21.3%, 16.1%, 31.8% and 25.0% respectively (Figure 4B). Notably, the incidence of severe treatment-related deterioration showed a clear upward trend with increasing score (6.0%, 12.0%, 12.9%, 31.8% and 25.0%). (Figure 4B).

**Figure 4.**
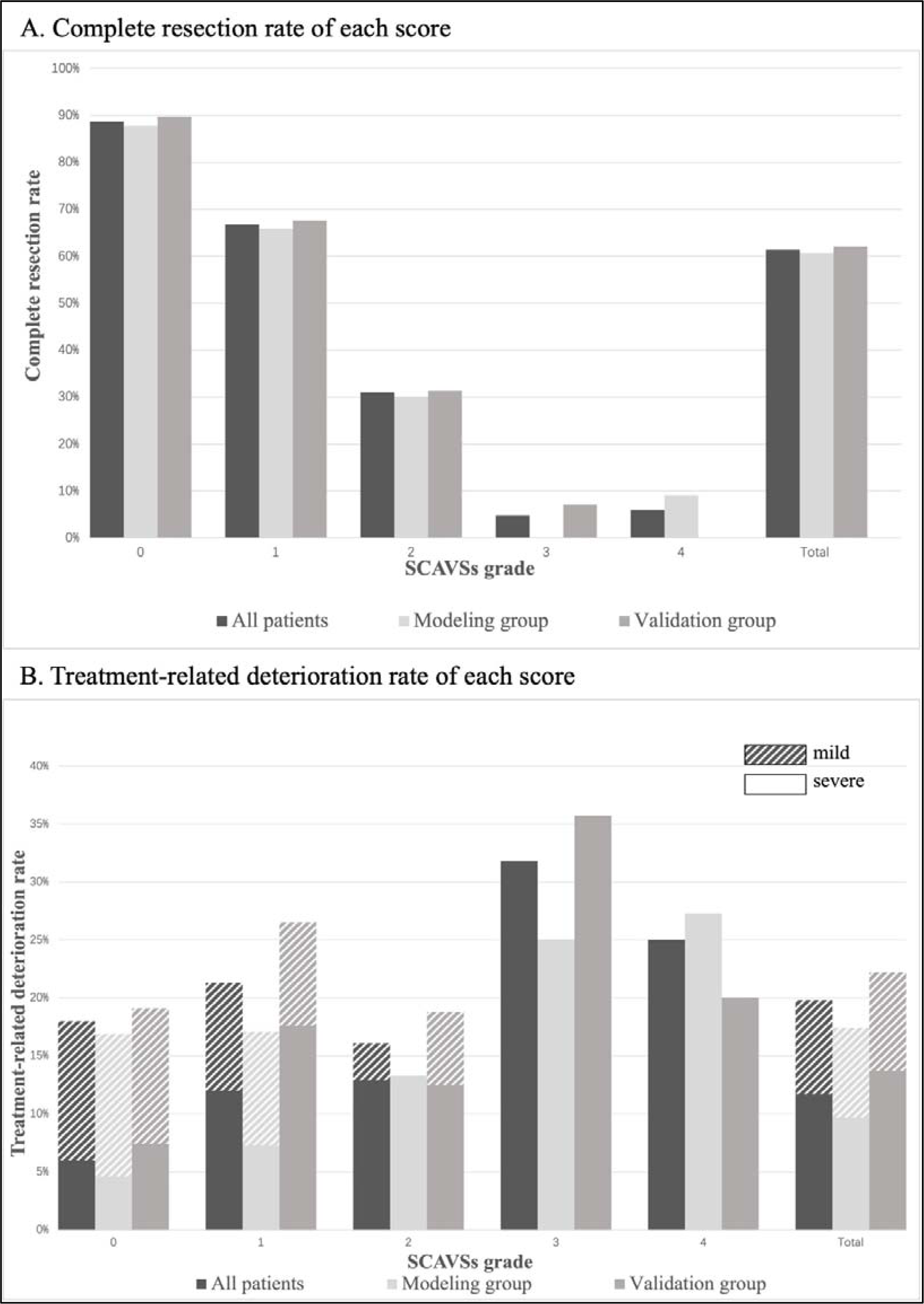
Complete resection rate and Treatment-related deterioration rate of each score. A: The complete obliteration rate declined as the score increased. B: An upward trend was observed in the incidence of treatment-related deterioration, especially the severe treatment-related deterioration, as the score increased.

Among patients with a score of 0, there was 36.0% of the treatment-related deterioration was classified as severe, which is significantly lower than that observed in other groups (77.8%), (p=0.001) (Figure 5).

**Figure 5.**
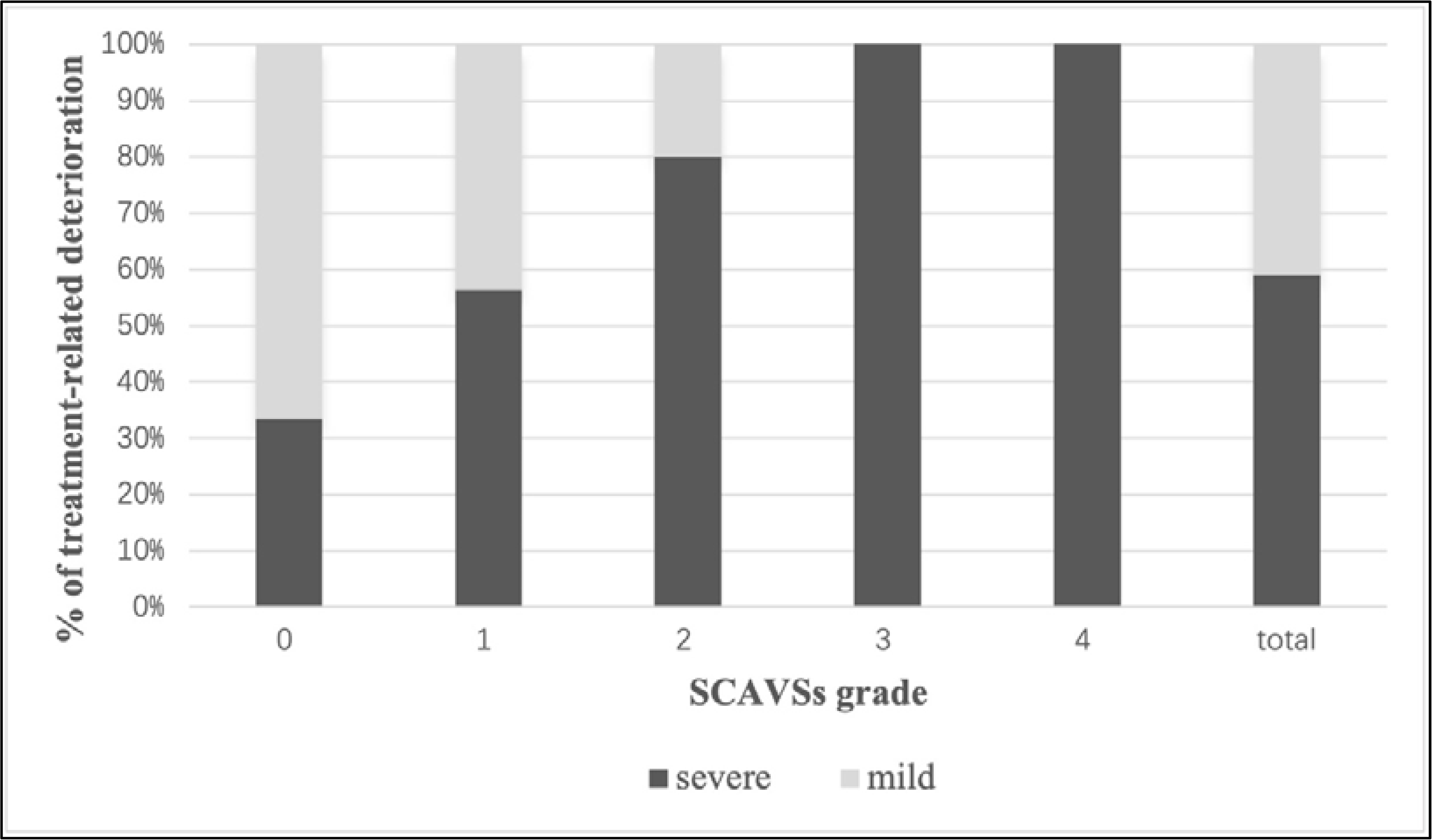
Severity of treatment-related deterioration of each score. The severity of treatment-related deterioration escalated as the score increased. Among patients with a score of 0, 36.0% of the treatment-related deterioration was classified as mild, which was significantly lower than that of other scores (77.8%), (p=0.001).

## Discussion

Based on the largest cohort of surgical managed SCAVSs, we proposed a concise surgical difficulty grading system that was determined by four parameters. This grading system could discernibly stratify the surgical feasibility for SCAVSs and was easy to apply for neurosurgeons specializing in spinal vascular diseases. The simplicity and universality of the scale rendered it a valuable tool for clinical decision-making and also be used as a reference point for evaluating treatment outcomes across different centers and surgical techniques.

The chosen parameters for the grading system are congruent with clinical practices. The anterior sulcal artery, as the primary branch of anterior spinal cord artery, radially supplies to the ventral 2/3 of the spinal cord from the ventral median fissure^17–18^. Conversely, the vast majority of SCAVSs surgeries were performed via posterior approach, which mean it is hard to control the blood supply originating from the ventral side of the cord during the initial phase of the operation^19^. Therefore, the anterior sulcal artery supply means we may encounter an irritable nidus with a higher risk of intraoperative bleeding as well as iatrogenic spinal cord impairment. The grading system also weighs the impact of lesion size and depth on surgical complexity. Undoubtedly, resection of an AVS with larger length and deeper location would pose greater challenge due to the inevitable disruption of more spinal cord itself and its blood supply^12,19^. Meanwhile, a large and embedded AVS usually receives blood supply from the anterior sulcal artery (Supplement Table 1).

The management of SMAVSs poses a great challenge due to the significantly more intricate angioarchitecture of these lesions compared to non-metameric counterparts^1–2,20–21^. Patients with SMAVSs are more prone to harboring a larger and deeper embedded nidus and usually fed by anterior sulcal artery (Supplement Table 2). However, it is noteworthy that even selected SMAVSs patients with small and superficial spinal cord AVS still achieved significantly lower complete resection rate compared to non-metameric AVSs under similar conditions (30.5% VS 69.1%, P<0.001). The primary factor contributing this discrepancy is that the intramedullary nidus of SMAVSs typically dispersed throughout the spinal cord tissue and may be insidious in the preoperative DSA (Figure 1). The insidious lesions are tiny AVSs that scattered in the spinal cord. When accompanied by extremely high-flow AVSs, there is no sufficient contrast agents for them to be detected until the high-flow AVSs are occluded. We hypothesize these phenomena may be associated with the etiology of the metameric manifestation of AVSs. Previous studies suggested that the pathogenic mutation of SMAVSs may occur prior to the migration of original vasculature to the surface of the spinal cord^1,22–24^. Therefore, the intramedullary vessels in patients with SMAVSs are bound to diffusely carry the pathogenic mutations which have the potential to form AVSs^4,25^. If the hypothesis were true, it may also suggest that the SMAVSs is associated with a higher risk of recurrence during long-term follow-up. However, due to the insufficient follow-up data, a definitive conclusion cannot be drawn currently.

In contrast to Spetzler-Martin scores^8^, the grading system for SAVSs stratifies the surgical feasibility based on both the probability of achieving complete resection and the treatment risk, rather than solely focusing on the risk of surgical-related deficits. Because the high eloquence of the entire spinal cord tissue, parenchymal resection, which is the most common procedure of BAVM resection, would be strictly prohibited^8,26–27^. The resection of SAVSs is mainly performed through sharp dissection at the interface between malformed vasculature and spinal cord tissue (Video 1). However, the extensive intramedullary exploration for complex nidus would still lead to neurological deficits^12,28^. Therefore, if intraoperative MEP or SEP falls below the threshold^14^, the procedure is usually terminated.

Based on the current analysis, we proposed the following recommendations for surgical decision-making. The data indicated that lesions scoring 0 points had a great chance of complete resection. Given the curative operation would eliminate the clinical risks of spinal cord impairment completely, it is therefore recommended that this subset of patients undergo microsurgery. The treatment-related risk of 18% appears to be significant when compared to other procedures within the field of neurosurgery. However, it should be noted that spinal cord AVSs have a poor natural disease course even when the lesion is partially obliterated^1–2,29^. Given that these diseases typically present in teenagers and young adults aged 20 to 30 years^1,30^, patients with residual lesions still face a grim prognosis due to the substantial cumulative risk of spinal cord impairment and enduring psychological distress throughout their lives^1–2^. Therefore, if microsurgery can effectively and promptly eliminate all clinical risks associated with the disease, it is reasonable to have a relatively higher tolerance for treatment-related risks. Additionally, considering that 64% of the treatment-related deterioration of patients with a score of 0 was classified as mild deficit, microsurgery should be an optimal choice for them. For patients with 2 to 4 points, our results showed a discouraging prospect for surgical intervention. Due to the intricate angioarchitecture and their intertwined intramedullary infiltration^12–14^, the operation was inevitably associated with considerable risks and incomplete resection. Thus, the surgical resection should not be considered as the first-line treatment option for this particular patient subset. We had determined that microsurgery may be considered as an option for patients with 1 point, however, the decision should be based on a meticulous evaluation of individual clinical risks, taking into account clinical features and angioarchitectural characteristics of the lesions. For high-risk patients (such as those with a history of recurrent spinal hemorrhage, signs of venous hypertension, aneurysmal structure or obvious mass effect)^1–2,20^, surgery aimed at removing high-risk components and reducing shunt flow should be considered instead of curative resection. Conversely, in patients with mild clinical risk and symptoms, microsurgery may not be suggested due to safety concerns.

## Limitation

Although the microsurgical techniques and the understanding of spinal cord vasculature anatomy have improved in the past decades, surgical management of SCAVSs remains a challenging task that requests experienced senior surgeons. Therefore, the representativeness of the treatment outcomes from our institutes should be validated by other specialized referral centers for spinal vascular diseases. Another limitation lies in the unsuitability of this grading system for anterior approach surgery. The vast majority of spinal vascular diseases surgery was performed by posterior approach^11^. However, the anterior approach surgery may be occasionally alternative for selected patients (e.g., superficial AVS ventrally located in the cervical cord), and our current data is insufficient to estimate the surgical feasibility for such cases.

## Conclusions

The proposed grading system, based on feeding artery origin, metameric manifestation, lesion size and depth, can effectively stratify the surgical difficulty of SCAVSs and assist in developing more rational treatment strategies. Based on this system, we recommend microsurgery for patients with 0 points, while surgical resection should not be used as the first-line treatment for patients with 2-4 points; for patients with 1 point, microsurgery is an optional approach aimed at removing high-risk components and reducing shunt flow, rather than achieving curative resection.

## Data Availability

Data are available upon reasonable request.

## Acknowledgements

We want to thank all the patients for their participation in this research.

## Author Contributions

Jia-Xing Yu, Hao-Han Lu, Li-Song Bian, Hong-Qi Zhang and Tao Hong contributed to the conception and design of the study; Chuan He, Fan Yang, Feng Ling, Jia-Xing Yu, Jian Ren, Jing-Wei Li, Hao-Han Lu, Hong-Qi Zhang, Gui-Lin Li, Li-Song Bian, Li-Yong Sun, Ming Ye, Peng Hu, Tao Hong, Yue-Shan Feng, and Yong-Jie Ma contributed to the acquisition and analysis of data; Jia-Xing Yu, Hao-Han Lu, Li-Song Bian and Yue-Shan Feng contributed to drafting the text or preparing the figures.

